# Scientific hypothesis generation process in clinical research: a secondary data analytic tool versus experience study protocol

**DOI:** 10.1101/2022.05.21.22275060

**Authors:** Xia Jing, Vimla L. Patel, James J. Cimino, Jay H. Shubrook, Yuchun Zhou, Chang Liu, Sonsoles De Lacalle

**Affiliations:** College of Behavioral, Social, and Health Sciences, Clemson University, Clemson, South Carolina, USA; Cognitive Studies in Medicine and Public Health, The New York Academy of Medicine, New York, New York, USA; Informatics Institute, School of Medicine, University of Alabama, Birmingham, Alabama, USA; College of Osteopathic Medicine, Touro University, Vallejo, California, USA; Patton College of Education, Ohio University, Athens, Ohio, USA; Russ College of Engineering and Technology, Ohio University, Athens, Ohio, USA; College of Art and Science, California State University Channel Islands, Camarillo, California, USA

**Keywords:** Clinical research, Observational study, Scientific hypothesis generation, Secondary data analytic tool, Think-aloud

## Abstract

**Background:** Scientific hypothesis generation is a critical step in scientific research that determines the direction and impact of any investigation. Despite its vital role, we have limited knowledge of the process itself, hindering our ability to address some critical questions.

**Objective:** To what extent can secondary data analytic tools facilitate scientific hypothesis generation during clinical research? Are the processes similar in developing clinical diagnoses during clinical practice and developing scientific hypotheses for clinical research projects? We explore the process of scientific hypothesis generation in the context of clinical research. The study is designed to compare the role of VIADS, our web-based interactive secondary data analysis tool, and the experience levels of study participants during their scientific hypothesis generation processes.

**Methods:** Inexperienced and experienced clinical researchers are recruited. In this 2×2 study design, all participants use the same data sets during scientific hypothesis-generation sessions, following pre-determined scripts. The inexperienced and experienced clinical researchers are randomly assigned into groups with and without using VIADS. The study sessions, screen activities, and audio recordings of participants are captured. Participants use the think-aloud protocol during the study sessions. After each study session, every participant is given a follow-up survey, with participants using VIADS completing an additional modified System Usability Scale (SUS) survey. A panel of clinical research experts will assess the scientific hypotheses generated based on pre-developed metrics. All data will be anonymized, transcribed, aggregated, and analyzed.

**Results:** This study is currently underway. Recruitment is ongoing via a brief online survey ^1^. The preliminary results show that study participants can generate a few to over a dozen scientific hypotheses during a 2-hour study session, regardless of whether they use VIADS or other analytic tools. A metric to assess scientific hypotheses within a clinical research context more accurately, comprehensively, and consistently has also been developed.

**Conclusion:** The scientific hypothesis-generation process is an advanced cognitive activity and a complex process. Clinical researchers can quickly generate initial scientific hypotheses based on data sets and prior experience based on our current results. However, refining these scientific hypotheses is much more time-consuming. To uncover the fundamental mechanisms of generating scientific hypotheses, we need breakthroughs that capture thinking processes more precisely.

## Introduction

A hypothesis is an educated guess or statement about the relationship between 2 or more variables ^2,3^. Scientific hypothesis generation is a critical step in scientific research that determines the direction and impact of research investigations. However, despite its vital role, we do not know the answers to some basic questions about the process. For example: can secondary data analytic tools facilitate the process? Is the scientific hypothesis generation process for clinical research questions similar to the differential diagnosis process? Traditionally, the scientific method involves delineating a research question and generating a scientific hypothesis. After formulating a scientific hypothesis, researchers design studies to test the scientific hypothesis to determine the answer to research questions ^2,4^.

Scientific hypothesis generation and scientific hypothesis testing are distinct processes ^2,5^. In clinical research, research questions are often delineated without the support of systematic data analysis (i.e., not data-driven) ^2,6,7^. Using and analyzing existing data to facilitate scientific hypothesis generation is considered ecological research ^8,9^. An ever-increasing amount of electronic data in health care are becoming available, much of which are coded. These data can be a rich source for secondary data analysis, accelerating scientific discoveries ^10^. Thus, many researchers have been exploring data-driven scientific hypothesis generation guided by secondary data analysis ^2,11^, including genomics ^5^. However, exactly how a scientific hypothesis is generated, even as shown by secondary data analysis in clinical research, is unknown. Understanding the detailed process of scientific hypothesis generation could improve the efficiency of delineating clinical research questions and, consequently, clinical research. Therefore, we are investigating the process of formulating scientific hypotheses guided by secondary data analysis. Using these results as a baseline, we plan to explore ways of supporting and improving the scientific hypothesis-generation process and study the process of formulating research questions as long-term goals.

Electronic health record (EHR) systems, and other technologies, are widely adopted in both office-based physician practices (86% in 2019 ^12^) and hospitals (overall 86% in 2022 and varies based on types of hospitals ^13^) across the United States. Thus, many electronic data are continuously captured and available for analysis to guide future decisions, uncover new patterns, or identify new paradigms in medicine. Much of the data is coded by hierarchical terminologies, such as the International Classification of Diseases Ninth Revision-Clinical Modification (ICD9-CM ^14^) and Tenth Revision-Clinical Modification (ICD10-CM ^15^), Systematized Nomenclature of Medicine-Clinical Terms (SNOMED-CT ^16^), Logical Observation Identifiers Names and Codes (LOINC ^17^), RxNorm ^18^, Gene Ontology (GO ^19^), and Medical Subject Headings (MeSH ^20^). We use the coded data sets by hierarchical terminologies ***as examples*** of existing data sets to facilitate and articulate the scientific hypothesis-generation process in clinical research, especially when guided by secondary data analysis. We developed algorithms ^21,22^ and a web-based secondary data analytic tool ^23-26^ to use the coded electronic data (ICD9 or MeSH) to conduct population and other clinically relevant studies.

Arocha and Patel ^27,28^ studied the directionality of reasoning (i.e., forward: from evidence to a scientific hypothesis, and backward: from a scientific hypothesis to evidence) in scientific hypothesis-generation processes and evaluation strategies (i.e., confirmation or disconfirmation) in solving a cardiovascular diagnostic problem by medical students (novice) and medical residents (experienced). More experienced clinicians used underlying situational knowledge about the clinical condition, while the novices used the surface structure of the patient information during the diagnostic generation process. Patel’s ^29,30^ and Kushniruk’s ^31^ studies used inexperienced and experienced clinicians with different roles, levels of medical expertise, and corresponding strategies to diagnose an endocrine disorder. In these studies, expert physicians used more efficient strategies (integrating patient history and experts’ prior knowledge early) to make diagnostic decisions ^29,31,32^. All these studies focused on hypothesis generation in solving diagnostic problems. Their results set the groundwork for reasoning in medical diagnostic process. Their findings (i.e., inexperienced and experienced clinicians generate diagnostic hypotheses via different processes) helped us formulate and narrow our research questions. Their methodology involved performing pre-defined tasks, recording “think-aloud” sessions, and transcribing and analyzing the study sessions. Making a diagnosis is a critical component of the medicine and a routine task for physicians. If generating the initial diagnostic hypothesis is correct, there is no need to search further for alternative hypotheses related to a particular patient.

On the other hand, generating scientific hypotheses in clinical research focuses on establishing a scientific hypothesis or doing further searches to explore alternative scientific hypotheses for research. The difference between the two can be demonstrated by the two enterprises, where the goal of generating a diagnostic hypothesis is to make decisions about a patient, and it is time constraint. In scientific research, time is not constrained, and the task is to explore various scientific hypotheses to formulate and refine the final research question. In both making a medical diagnosis in clinical practice and scientific thinking, generating initial diagnostic hypotheses depends on prior knowledge and experience ^33,34^. However, in scientific thinking, analogy and associations play significant roles in addition to prior knowledge, experience, and reasoning capability. Analogies are widely recognized as playing a vital heuristic role as aids to discovery ^33^. They have been employed in a wide variety of settings and had considerable success in generating insight and formulating possible solutions to existing problems.

This makes it essential for us to understand the scientific hypotheses generation process in clinical research and compare this process to generating a clinical diagnosis, including the role of experience during the processes.

The current study explores the scientific hypothesis-generation process in clinical research. This study investigates whether a secondary data analytic tool and clinician experience affect the scientific hypothesis-generation process. We propose to use direct observations, think-aloud methods with video capture, follow-up inquiry and interview questions, and surveys to capture the participants’ perception of the scientific hypothesis-generation process and associated factors. The qualitative data generated will be transcribed, analyzed, and quantified.

In the study, we aim to test the following study hypotheses.

- Experienced and inexperienced clinical researchers will differ in generating scientific hypotheses guided by secondary data analysis.
- Clinical researchers will generate different scientific hypotheses with and without using a secondary data analytic tool.
- Researchers’ level of experience and use of secondary data analytic tools will interact in their scientific hypothesis-generation process

In this paper, we use the term “research hypothesis” to refer to a statement generated by our research participants, the term “study hypothesis” to refer to the subject of our current research study, and the term “scientific hypothesis” to refer to the general mention of hypotheses in research contexts.

## Methods

We use a mixed-method study design. The study includes direct observational, utility, and usability studies. Surveys, interviews, semi-predefined tasks, and capturing screen activities are all used. The modified Delphi method is also used in the study.

### Participants and Recruitment

Experienced and inexperienced clinical researchers and a panel of clinical research experts are recruited. The main criteria to separate the three groups are based on their experience in clinical research. Table 1 summarizes the requirements for clinical researchers, expert panel members, and the computers that clinical researchers use during the study sessions. Participants are compensated for their time according to the professional organizations’ guidelines. The study has been approved by the Institutional Review Board (IRB) at Clemson University (IRB2020-056).

**Table 1.**
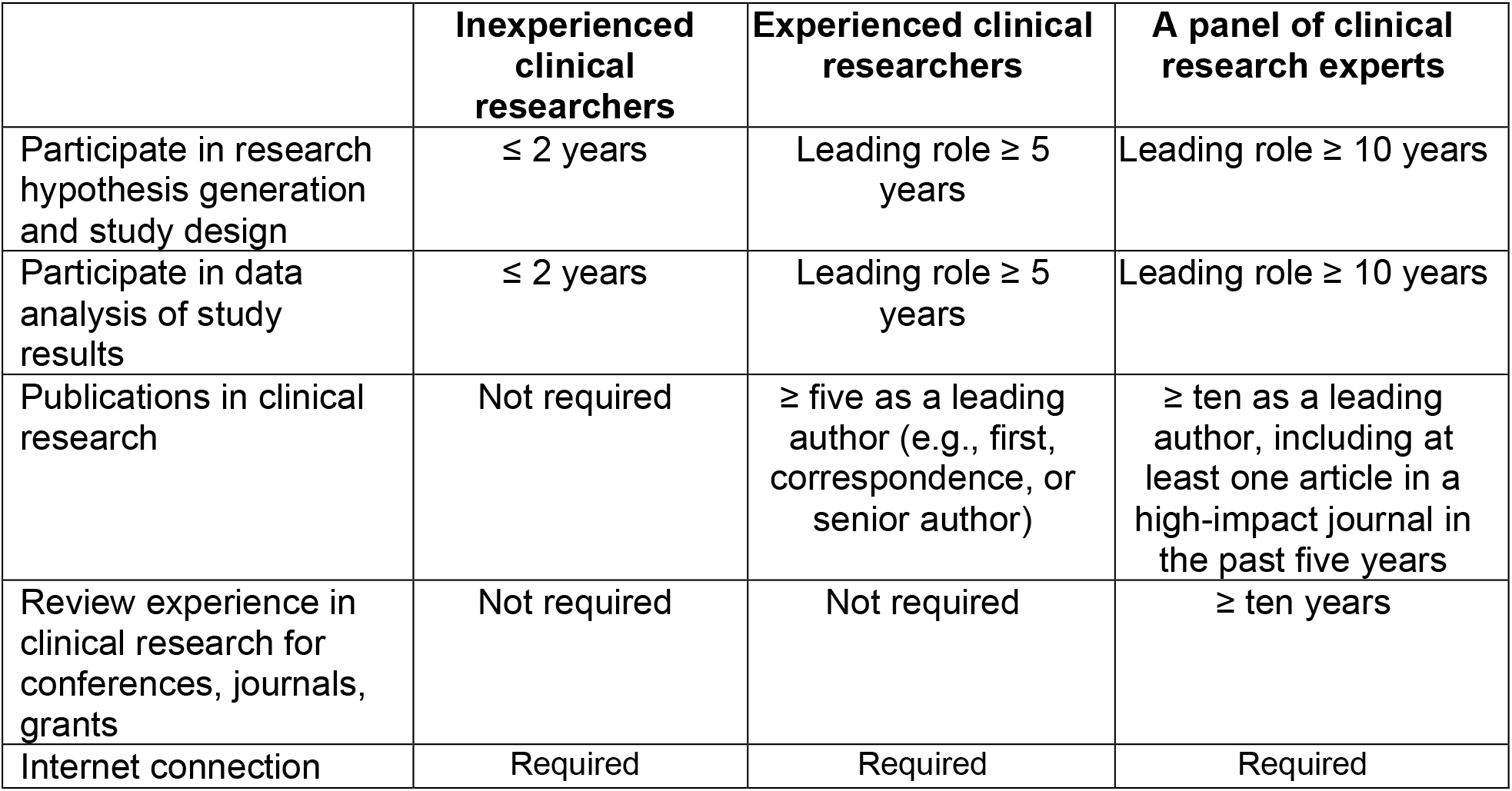
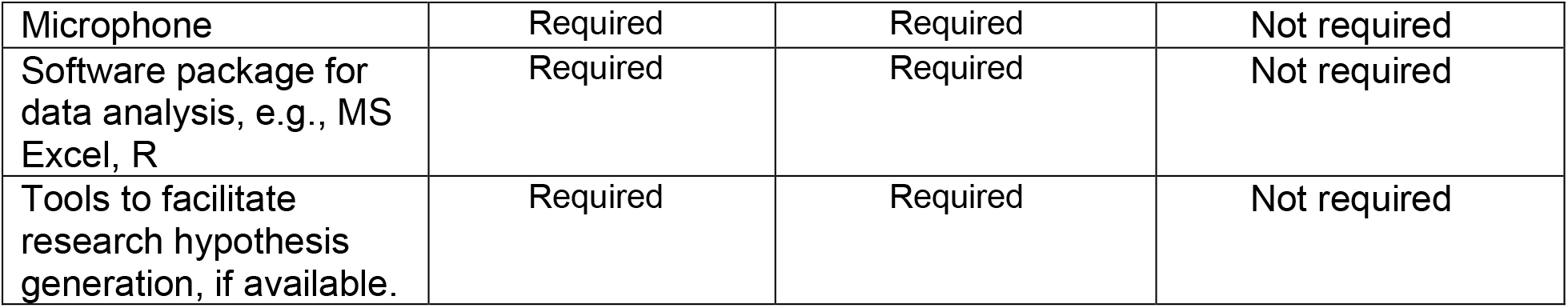
Summary criteria for study participants and clinical research expert panel members

To recruit participants, invitational emails and flyers were sent to collaborators, and through other communications, such as mailing lists (e.g., working groups of the American Medical Informatics Association), newsletters (e.g., South Carolina Clinical & Translational Research Institute newsletters and PRISMA health research newsletters), and Slack channels (e.g., National COVID Cohort Collaborative communities). All study sessions are conducted remotely via video conference software.

### Introduction to VIADS

In this study, we use VIADS as an example of a secondary data analytic tool. VIADS is a visual interactive analytical tool for filtering and summarizing large health data sets coded with hierarchical terminologies ^23,24,35^. It is a cost-free Web-based tool available for research and educational purposes. VIADS can be used as registered users or as guest users without registration. Our group developed VIADS as well as the underlying algorithms over the years ^22,25,36^. VIADS is designed to use codes from terminologies with hierarchical structures (for example, ICD9-CM, MeSH) and their usage frequency to achieve the following objectives: (1) provide summary visualization (i.e., graphs) of a dataset; (2) filter data sets to ensure manageable sizes based on users’ selection of algorithms and thresholds; (3) compare similar datasets and highlight the differences; (4) provide interactive, customizable, and downloadable features for the graphs generated from the datasets. VIADS is a useful secondary data analytic tool that can facilitate medical administrators, clinicians, and clinical researchers make their respective decisions. For example, VIADS can be used to track longitudinal data of a hospital over time and explore the trends and detect the differences in diagnoses over time; VIADS can also be used to compare two similar medications and compare the medical events associated with the medications to provide detailed evidence to guide more precise clinical use of the medications ^36^. Our study provided evidence of the different information needs between physicians and nurses via the algorithms of VIADS ^22^. Figure 1 shows example screenshots of VIADS.

**Figure 1.**
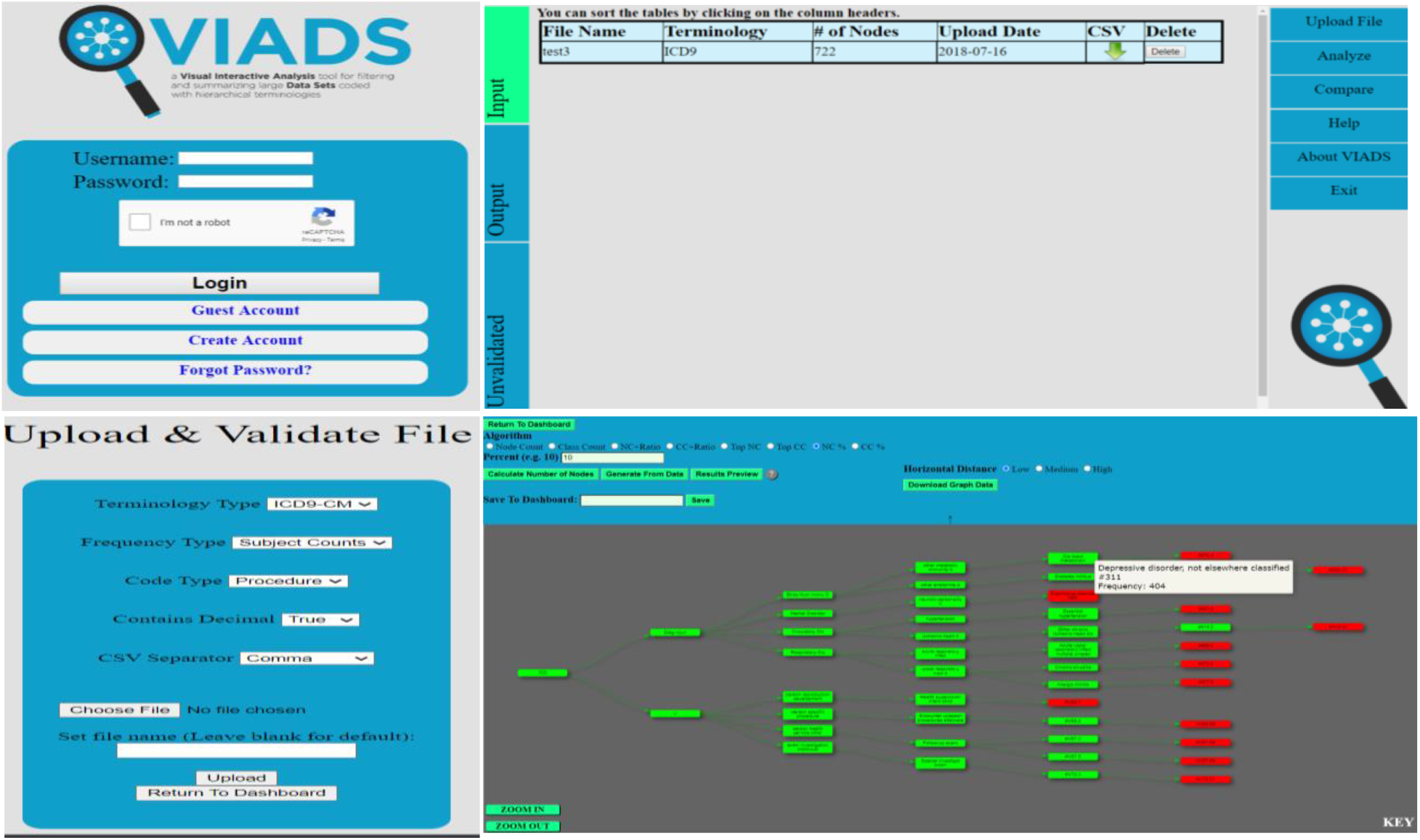
Selected screenshots of VIADS (homepage—upper left; validation module—lower left; dashboard—upper right; a graph generated—lower right)

Meanwhile, we recognize that VIADS cannot accept any types of clinical data but coded data and their use frequencies. This can limit the types of scientific hypotheses generated while using VIADS.

### Preparation of Test Data Sets

We prepare and use the same data sets for this study across different groups to reduce the potential bias introduced by different data sets. However, all data sets (i.e., input files) used in VIADS are usually prepared by users in individual institutions. Table 2 summarizes the final format of data sets and the minimum accepted sizes of data sets needed to analyze in VIADS. The current version of VIADS is designed to accept all data sets coded by the three types of terminologies (ICD9, ICD10, and MeSH) listed in Table 2. No identified patient information is included in the data sets used in VIADS, as the data sets contain only the node identification (i.e., terminology code) and usage frequencies.

**Table 2.** Acceptable format and sizes of data sets in VIADS

|  | **Graph node ID (code)** | **Usage frequency** |
| --- | --- | --- |
| **ICD9-CM** data set | 300.00 | 2223 |
|  | 278.00 | 5567 |
|  | ... .. | ... |
| **ICD10-CM** data set | O10.01 | 5590 |
|  | E11.9 | 50000 |
|  | ... .. | ... |
| **MeSH** data set | A0087342 | 16460 |
|  | A0021563 | 4459 |
|  | ... .. | ... |
| Acceptable data set size for Web VIADS | **Patient counts** | ≥ 100 |
|  | **Event counts** | ≥ 1000 |

Abbreviations: ICD9-CM, International Classification of Diseases Ninth Revision-Clinical Modification; ICD10-CM, International Classification of Diseases Tenth Revision-Clinical Modification; MeSH, Medical Subject Headings.

The usage frequencies of the data sets used in VIADS can be:

- Patient counts: The number of patients associated with ICD codes in the selected database.
- Event counts: The number of events (ICD codes or MeSH terms) in the selected database.

An ancestor-descendant table, which contains one row for each node and each of its distinct descendants, can calculate class counts easier and more accurately without counting the same node multiple times. Such implementation details are discussed in our prior publications ^21,22^.

We use a publicly accessible data source^37^, including ICD9-CM codes, to generate the needed input data sets. We use the patient counts as frequencies. Although ICD10-CM codes are now used in the United States, ICD9-CM data from the past several decades are available in most institutions across the country. We use ICD9-CM codes to obtain historical and longitudinal perspectives.

### Instrument Development

We developed metrics to assess research hypotheses generated during the study sessions. The development process goes through iterative stages (Figure 2) via Qualtrics surveys, emails, and phone calls. First, we conducted the literature review to outline the draft metrics. Then we discussed the draft metrics and iteratively revised them until all concerns were addressed. Then the revised metrics were distributed to the entire research team for feedback. The internal consensus processes follow a modified Delphi method ^38^, including transparent and open discussions (via email replies) among the entire research team and anonymous survey responses.

**Figure 2.**
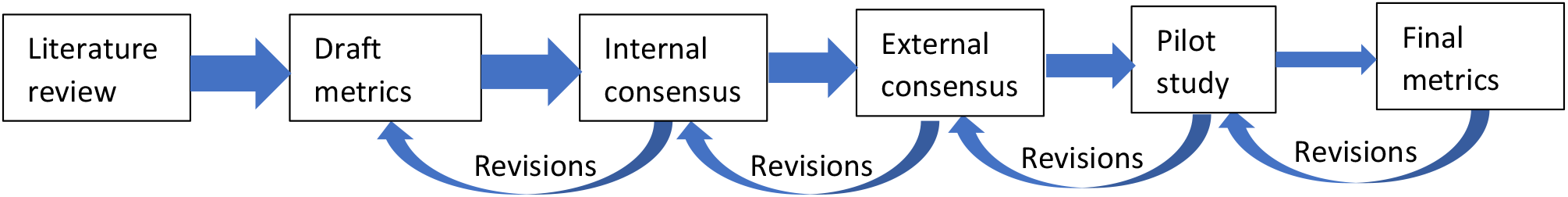
Development process for metrics to evaluate research hypothesis in clinical research.

The performance of scientific hypothesis-generation tasks will be measured by metrics that include qualitative and quantitative measures: **validity, significance, clinical relevance, feasibility, clarity, testability, ethicality**, number of total scientific hypotheses, and average time used to generate one scientific hypothesis. In an online survey, the panel of clinical research experts will assess the generated research hypotheses based on the metrics we developed.

A survey (Appendix A) at the end of the research hypothesis-generation study sessions is administered for the first four groups (Figure 3). The groups that use VIADS also complete a modified SUS questionnaire (Appendix B) about the usability and utility of VIADS. The follow-up or inquiry questions during the think-aloud process used in Study 1 (Figure 3) are as follows:

- What are needed but not available attributes to help clinical researchers generate research hypotheses (i.e., a wish list of features facilitating research hypothesis generation)?
- Follow-up questions to clarify potential confusion during the think-aloud processes or conduct a meaningful inquiry when unexpected or novel questions emerge during observations ^39^. However, these questions are kept at a minimal level to avoid interrupting clinical researchers’ thinking processes. This method complements the data from the think-aloud video captures.
- Whether the list of items in Appendix C, which is from the traditional clinical research textbooks ^2-4,9,40^ on scientific hypothesis-generating and research-question formulating, can facilitate clinical researchers’ research hypothesis generation when guided by secondary data analysis.

**Figure 3.**
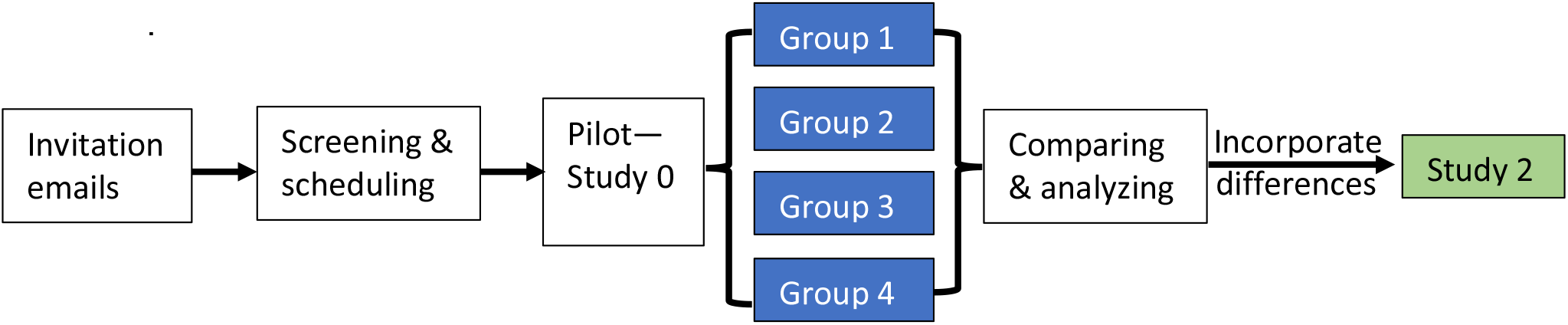
Summary of the study procedures (blue boxes indicate data collected in Study 1; please refer to Table 3 for the details of different groups)

A survey will be developed based on the comparative results of the first four groups and administered at the end of Study 2 (i.e., the identified differences from Study 1 will be the focus of the survey to determine whether these differences are helpful in Study 2).

### Study Design

Study 1 tests all three study hypotheses. Suppose the study hypotheses are supported in Study 1. In that case, we will conduct a follow-up study (Study 2) to examine whether and how the efficiency or quality of research hypothesis generation may be improved or refined.

We use the think-aloud protocol method to conduct the following tasks: (1) observe the research hypothesis-generation process and (2) transcribe data, analyze and assess if VIADS^23^ and different levels of experience of clinical researchers influence the process, and (3) assess the interactions between VIADS (as an example of secondary analytic tools) and experience levels of the participants during the processes. Figure 3 summarizes the Study 1 and Study 2 procedures.

#### Study 1

Experienced and inexperienced clinical researchers conduct research hypothesis-generation tasks using the same secondary datasets. We capture the tasks to describe participants’ research hypothesis-generation process when guided by analysis of the same datasets. We also use VIADS as an example of a secondary data analytic tool to assess participants’ research hypothesis-generation process. Accordingly, the two control groups do not use VIADS, but the two intervention groups do.

A pilot study (i.e., Study 0) was conducted for each group for the feasibility of using the task flow, datasets, screen capture, audio and video recordings, and the study scripts. In study 1, we use four arms. Table 3 summarizes the study design and participants in each arm. The four arms will be compared to detect the main effects of the two factors and their interactions after completing Study 1. After recruiting, the participants are randomly assigned to groups using VIADS (3-hour session, 1 hour to conduct VIADS training) or without using VIADS (2-hour session).

**Table 3.** Design of study 1 for research hypothesis-generation process in clinical research

|  | **# of Experienced clinical researchers** | **# of Inexperienced clinical researchers** |
| --- | --- | --- |
| **Without using VIADS** | **8 (Group 1)** | **8 (Group 2)** |
| **Using VIADS** | **8 (Group 3)** | **8 (Group 4)** |

We conduct one study session with each participant individually. Each participant is given the same datasets to generate research hypotheses during each study session with or without VIADS. The same researcher observes the entire process and captures the process via the recording of think-aloud videos. Follow-up or inquiry questions from the researcher (i.e., observer) are used as complementary during each study session. The observation is conducted remotely via Webex. All screen activities are captured and recorded via BB FlashBack ^41^.

#### Study 2

If the results of Study 1 indicate group differences (i.e., there are differences between experienced and inexperienced clinical researchers; and there are differences between groups using VIADS and without using VIADS) as we expect, then we will conduct Study 2 to examine whether the efficiency of the research hypothesis-generation process could be improved. Specifically, we will analyze for group differences, identify anything related to VIADS and incorporate them into VIADS. Then, we will test whether the revised (i.e., improved) VIADS can increase the efficiency or quality of the research hypothesis-generation process. In this approach, the group that performs less well in Study 1 will be invited to use the revised VIADS to conduct research hypothesis-generation tasks again with the same datasets. However, at least eight months apart to ensure an adequate wash-out period. Their performance will be compared with that in Study 1.

Suppose no significant difference can be detected between the groups using VIADS and without using VIADS in Study 1. We will use the usability and utility survey results to revise VIADS accordingly without conducting Study 2. If no significant difference can be detected between experienced and inexperienced clinical researchers, then Study 2 will recruit both experienced and inexperienced clinical researchers as study participants. In this case, Study 2 will focus only on whether revised VIADS impacts the research hypothesis-generation processes and outcomes.

### Data and Statistical Analysis

While conducting the given tasks, the qualitative data collected via the think-aloud method will be transcribed, coded, and analyzed according to the grounded theory ^42,43^, a classical qualitative data analytic method. The data analysis has not started yet since we are still collecting data. Combination analysis of discourse ^5^, video (i.e., study sessions), and screen activities will be conducted. The main components or patterns that we will focus on during analysis include potential nonverbal steps, the sequential ordering among different components (such as using experience or data first) across groups, seeking and processing evidence, analyzing data, generating inferences, making connections, formulating a hypothesis, the searching for information needed to generate research hypotheses, and so forth. Ideally, based on the video analysis and observations, we plan to develop a framework for the scientific hypothesis-generation process in clinical research guided by secondary data analysis. Similar frameworks exist in education and learning areas ^44^ but are not currently in clinical research.

The outcome variable is participants’ performance in research hypothesis-generation tasks. The performance is measured by the quality (e.g., significance, validity) and quantity of the research hypotheses generated through the tasks and the average time to generate one research hypothesis. At least three clinical research experts will assess each hypothesis using the scientific hypothesis assessment metrics we developed for this study. The metrics include multiple items, each on a 5-point scale. The details of the metrics are described in the section on Instrument Development.

The data will be analyzed with a two-tailed factorial analysis. We calculated the required sample size as 32 based on a confidence level of 95% (α = 0.05) and a power level of 0.8 (β = 0.20). A *t*-test will be conducted to determine whether the revised VIADS improves the performance of research hypothesis generation in Study 2.

## Results

The study is a National Institutes of Health-funded R15 project supported by the National Library of Medicine. This study is currently underway, and herein we share some preliminary results and summarize our early observations to date. We will share the full results and analysis of the study in future publications.

### Instruments

Based on our literature review, we developed metrics to assess research hypotheses ^2,3,5,7-9,40,45^. Most of the dimensions used to evaluate clinical research hypotheses include clinical and scientific ***validity, significance*** (regarding the target population, cost, future impact), ***novelty*** (regarding new knowledge, impact on practice, new methodology), ***clinical relevance*** (regarding medical knowledge, clinical practice, policies), ***potential benefits and risks, ethicality, feasibility*** (regarding cost, time, the scope of the work), ***testability, clarity*** (regarding purpose, focused groups, variables, their relationships), and ***researcher interest level*** (i.e., willingness to pursue). Multiple items were used to measure the quality for each dimension mentioned above. Under each item, we use a 5-point Likert scale (1 = strongly disagree, 5 = strongly agree) to measure it. After internal consensus, we conduct external consensus and seek feedback from the external expert panel via an online survey ^46^. The metrics are revised continuously by incorporating feedback. The expert panel will use our online survey ^47^ to evaluate research hypotheses generated by research participants during the study sessions.

We developed the initial study scripts for Study 1 and revised them after the pilot study sessions (Study 0). We developed the screening survey for the recruitment process. We also developed the follow-up survey after each study session, regardless of the groups. We modified the standard SUS survey ^48,49^ to add one more option to let users elaborate on what caused their dissatisfaction during the usability study, if there is any.

### Recruitment

We are recruiting all participants: inexperienced clinical researchers, experienced clinical researchers, and a panel of clinical research experts. To participate, anyone in clinical research can share their contact email address by filling out the screening survey ^1^. We have completed 12 study sessions with inexperienced clinical researchers using or not using VIADS in Study 1.

### Study 1

We use data from the National Ambulatory Medical Care Survey (NAMCS) conducted by the Centers for Disease Control and Prevention ^37^. The NAMCS is a publicly accessible dataset of survey results related to clinical encounters in ambulatory settings. We processed raw NAMCS data (ICD9 codes and accumulated frequencies) in 2005 and 2015 to prepare the needed datasets for VIADS based on our requirements.

To determine which group a participant joins for both inexperienced (groups 2 and 4) and experienced (groups 1 and 3) clinical researchers, we used an R package (blockrand ^50^) to implement block randomization. The random blocks range from 2 to 6 participants.

## Discussion

A critical first step in the life cycle of any scientific research study is formulating a valid and significant research question, which usually can be split into several scientific hypotheses. This process is often challenging and time-consuming ^2,4,39,51^. Currently, there is limited practical guidance regarding generating research questions ^39^ beyond emphasizing that it requires long-term experience, observation, discussion, and exploration of the literature. Scientific hypothesis generation will eventually help to formulate relevant research questions. Our study aims to decipher the process of scientific hypothesis generation and determine whether a secondary data analytic tool can facilitate the process in a clinical research context. When combined with clinical researchers’ experience and observations, we anticipate that such tools will facilitate scientific hypothesis generation. This facilitation will improve the efficiency and accuracy of testing scientific hypotheses, formulating research questions, and conducting clinical research in general. We also anticipate that an explicit description of the scientific hypothesis-generation process with secondary data analysis may provide more feasible guidance for clinical research design newcomers (e.g., medical students and new clinical investigators). However, we have not completed all study sessions, so we cannot yet carefully analyze data to draw conclusions.

We observed that participants used analogical reasoning ^52^ consciously and subconsciously; meanwhile, some verbally expressed that they avoided analogical reasoning intentionally to be more creative (i.e., noting using the same pattern of statements for all the topics supported by the datasets) during the study sessions. The way we organized the datasets seems to promote the participants (i.e., users of datasets) to think systematically, e.g., the use frequencies of ICD9 codes were sorted from high to low in each dataset. However, what is the perfect balance between systematic structure and randomness during scientific hypothesis generation is unknown. Intuitively, both systematic reviewing and random connections should be critical in generating novel ideas in general, regardless of academic settings or industry environments. Concrete evidence is needed to draw any conclusions about the relationships between the two during scientific hypothesis generation with certainty.

Analyzing the research hypothesis-generation process may include several initial cognitive components. These components can consist of searching for, obtaining, compiling, and processing evidence; seeking help to analyze data; developing inferences using obtained evidence and prior knowledge; searching for external evidence (e.g., literature, prior notes), looking for connections between evidence and problems, considering feasibility (or testability, ethicality, and clarity), drawing conclusions, formulating draft research hypotheses, and polishing draft research hypotheses ^2-4,9,40,53^. These initial components will be used to code the recorded think-aloud sessions to compare differences among groups.

Establishing the evaluation metrics to assess research hypotheses is the first step and the critical foundation for the overall study. The evaluation metrics will determine the quality measurements of research hypotheses generated by the study participants during study sessions.

Research hypothesis evaluation is subjective, but metrics can help standardize the process to some extent. Although the metrics may not guarantee a precise or perfectly objective evaluation of each research hypothesis, such metrics provide a consistent instrument for this highly sophisticated cognitive process. We anticipate that the consistent instrument will help to standardize the expert panel’s evaluation. In addition to the metrics developed in this study, we will also use objective measures, such as the number of research hypotheses generated by the study participants and the average time each participant spends generating each research hypothesis. We expect the expert panel to provide more consistent research hypothesis evaluations with the combined metrics and objective measures.

Although developing metrics appears linear, as presented in Figure 2, the process itself is highly iterative. No revision occurs only once. When we reflect on the first three stages of development, we see that major revisions during the first three stages involve separating questions in the survey and refining the options for the questions. These steps reduce ambiguity.

### Challenges

We have faced many challenges conducting the research hypothesis-generation study sessions. These include: (1) What can be considered a research hypothesis? What will not be considered a research hypothesis? The response will determine which research hypotheses will be evaluated by the panel of clinical research experts. (2) How to measure a research hypothesis accurately? Although we developed our workable metrics, the metrics are not perfected, as yet. (3) How can we accurately capture thinking, reasoning, and networking processes during the research hypothesis-generation processes? Currently, we use the think-aloud method. Although think-aloud protocols can capture valuable information about the thinking process, we recognize that not all processes can be articulated during the experiments, and not everyone can articulate the process accurately. (4) What happens when a clinical researcher examines and analyzes a data set and generates a research hypothesis subconsciously. (5) How can we capture the roles of the external environment, internal cognitive capacity, existing knowledge body of the participant, and interactions between the individual with the external world play in these dynamics?

When faced with challenges, we see opportunities for researchers to explore further and identify a clearer picture of research hypothesis generation. We believe that the most pressing target is developing new technologies to capture what clinical researchers do and think when generating research hypotheses with a data set. Such technologies can promote breakthroughs in cognition, psychology, computer science, artificial intelligence, neurology, and clinical research in general. In clinical research, such technologies can help in empowering clinical researchers to conduct their tasks more efficiently and effectively.

### Initial Observations

We noticed that both forward ^28^ and backward reasoning were used during the study sessions. However, backward reasoning did not start from a scientific hypothesis in several study sessions. Instead, it started from the participant’s focused (and often familiar) area (i.e., participants’ focused on several ICD9 codes in the focused area). The research hypotheses were developed after looking into the data on the focused area.

Many participants did not use any advanced analysis during the study session. However, they used their prior experience and knowledge to generate research hypotheses based on the frequency rank of the provided datasets and by comparing the two years of data, i.e., 2005 versus 2015.

Noticeably, our VIADS can answer more complicated questions systematically and timely. However, we noticed that the training session of VIADS increased participants’ cognitive load. Without a comprehensive analysis, we cannot yet draw further conclusions.

We recognize that research hypothesis generation and the long refining and improving process matter most during the study sessions. Without technologies to capture what happens cognitively during the research hypothesis-generation process, we may not be able to answer the fundamental question about the mechanisms of scientific hypothesis generation.

### Lessons Learned so far

We learned some important lessons while designing and conducting our study. The first lesson was about balancing the study design (straightforward or complicated) and the study conduction (feasibility). During the design stage, we were concerned that the 2×2 study was too simple, even though we know it does not negatively affect the value of the research. We only considered experience levels and whether use VIADS for such a complicated cognitive process. However, even for such a straightforward design, the experienced clinical researchers only have one volunteer during recruitment thus far. Thus, we first focused on inexperienced clinical researchers. Even for inexperienced clinical researchers’ study sessions, we need considerable time to determine strategies for coding and analyzing the raw data. To design a complicated experiment that answers more complex questions, we must consider the balancing practical workload, recruitment reality, expected timeline, and researchers’ desire to pursue a complex research question.

Recruitment is always challenging. Many of our panel invitations to clinical research experts either received no response or were rejections, which significantly delayed the study timeline, in addition to the Pandemic. Further, the IRB approval process was time-consuming, delaying our study when we needed to revise study documents. The study timeline includes the IRB initial review and re-review cycles.

### Future Work

The first future direction for this project is to explore the feasibility of formulating research questions based on research hypotheses. In the current project, we look for ways to improve the efficiency of generating research hypotheses. The next step will be to explore whether we can enhance the efficiency of formulating research questions.

The second possible direction for future work is to develop tools to facilitate scientific hypothesis generation guided by secondary data analysis. We may explore automating the process or incorporating all positive attributes to guide the process better and improve efficiency and quality.

At the end of our experiments, we ask clinical researchers what facilitates their scientific hypothesis generation. Several of their responses included repeatedly reading and discussing with colleagues. We believe intelligent tools can undoubtedly improve both aspects of scientific hypothesis generation: summarizing new publications of the chosen topic areas and providing conversational support to clinical researchers. This would be a natural extension of our studies.

## Data Availability

This manuscript is the study protocol. After we analyze and publish the results, transcribed, aggregated, de-identified data can be requested from the authors.

## Acknowledgments

The project is supported by a grant from the National Library of Medicine of the United States National Institutes of Health (R15LM012941) and partially supported by the National Institute of General Medical Sciences of the National Institutes of Health (P20 GM121342). The content is solely the author’s responsibility and does not necessarily represent the official views of the National Institutes of Health.

## Conflicts of Interest

None to disclose.

## Abbreviations

GO: the Gene Ontology
ICD: the International Classification of Diseases
ICD9-CM: International Classification of Diseases Ninth Revision-Clinical Modification
ICD10-CM: International Classification of Diseases Tenth Revision-Clinical Modification
IRB: Institutional Review Board
LOINC: Logical Observation Identifiers Names and Code
MeSH: Medical Subject Headings
NAMCS: National Ambulatory Medical Care Survey
SNOMED-CT: Systematized Nomenclature of Medicine-Clinical Terms
SUS: System Usability Scale
VIADS: A visual interactive analysis tool for filtering and summarizing large data sets coded with hierarchical terminologies

## Appendices

## Appendix A

Follow-up survey at the end of the hypothesis-generation tasks

## Appendix B

VIADS SUS and utility questionnaire

## Appendix C

List of items to consider during hypothesis generation

## Notes

### Competing Interest Statement

The authors have declared no competing interest.

### Clinical Trial

This study is not a clinical trial per NIH definition.

### Author Declarations

The study has been approved by the Institutional Review Board (IRB) at Clemson University (IRB2020-056).

