## Appendix C for "Scientific hypothesis generation process in clinical research: a secondary data analytic tool versus experience study protocol"

### Appendix C: Dimensions to consider during the hypothesis generation

The existing literature provides some dimensions that should be considered during the hypothesis generation process for a research purpose; however, they are not specifically utilized for this study, i.e., secondary data analysis to guide hypothesis generation in clinical research. The following dimensions will be used as a checklist for study participants as a traditional way to facilitate their hypothesis generation. These dimensions include:

- Basic, applied, translational research
- Observational or experimental study
- Descriptive or analytic research
- Comparison, association, or causal study
- Retrospective or prospective research
- Longitudinal or cross-sectional research
- Qualitative or quantitative research
- Single variable or multiple variables
- Variables: dependent, independent, moderator, control, and intervening
- Focused population
- One-tailed or two-tailed
- Investigational intervention
- Outcomes of interest

These dimensions have been included in the study scripts for participants to reference during the study sessions.
