## Appendix B for "Scientific hypothesis generation process in clinical research: a secondary data analytic tool versus experience study protocol"

### VIADS system usability scale (SUS) and utility questionnaire

1. I would like to use VIADS frequently.
    A. Strongly disagree
    B. Disagree
    C. Neutral
    D. Agree
    E. Strongly agree

2. I find VIADS unnecessarily complex.
    A. Strongly disagree
    B. Disagree
    C. Neutral
    D. Agree
    E. Strongly agree
    - Please specify what you would do to make VIADS simpler if your answer is D or E for question 2. Please list three things you would change in VIADS.

3. I think VIADS is easy to use.
    A. Strongly disagree
    B. Disagree
    C. Neutral
    D. Agree
    E. Strongly agree
    - Please specify your answer or give an example in which VIADS is not easy to use if your answer is A or B for question 3.

4. I would need technical support in order to use VIADS.
    A. Strongly disagree
    B. Disagree
    C. Neutral
    D. Agree
    E. Strongly agree
    - Please specify what support (such as navigation, definitions of the terms used in VIADS interfaces, mechanism of VIADS behind the scenes etc.) you need to be able to use VIADS if your answer is D or E for question 4.

5. I find the various functions in VIADS are well integrated with each other.
    A. Strongly disagree
    B. Disagree
    C. Neutral
    D. Agree
    E. Strongly agree
    - Please specify your answer if your answer is A or B for question 5. Please give an example of how the various functions in VIADS are not well integrated.
    - Do you have any additional suggestions to make them better integrated?

6. I think there are too many inconsistencies in VIADS.
    A. Strongly disagree
    B. Disagree
    C. Neutral
    D. Agree
    E. Strongly agree
    - Please give examples of inconsistencies in VIADS if your answer is D or E for question 6.

7. I imagine that most people can learn to use VIADS quickly.
    A. Strongly disagree
    B. Disagree
    C. Neutral
    D. Agree
    E. Strongly agree
    - Please explain why you think most people cannot learn to use VIADS quickly if your answer is A or B for question 7.

8. I find VIADS cumbersome to use.
    A. Strongly disagree
    B. Disagree
    C. Neutral
    D. Agree
    E. Strongly agree
    - Please give examples that VIADS is cumbersome to use if your answer is D or E for question 8.

9. I feel confident using VIADS.
    A. Strongly disagree
    B. Disagree
    C. Neutral
    D. Agree
    E. Strongly agree
    - Please explain why you are not confident using VIADS if your answer is A or B for question 9. Please include examples.

10. I need to learn more before I can use VIADS.
    A. Strongly disagree
    B. Disagree
    C. Neutral
    D. Agree
    E. Strongly agree
    - Please specify what you need to learn before you can use VIADS if your answer is D or E for question 10. Please include examples.

11. Does VIADS provide new perspectives or measurements for understanding the data set?
    A. Yes
        i. If yes, please provide specifics___________
    B. No
    C. Other, please specify___________________

12. Do you think that VIADS can facilitate the interpretation of the data set?
    A. Yes
        i. If yes, please provide specifics___________
    B. No
    C. Other, please specify___________________

13. Do you think that VIADS can facilitate your decision making in generating hypothesis?
    A. Yes
        i. If yes, please provide specifics___________
    B. No
    C. Other, please specify___________________

14. Do you think that VIADS can facilitate your capability to present the data set?
    A. Yes
        i. If yes, please provide specifics___________
    B. No
    C. Other, please specify___________________

15. Do you think that VIADS can be used to answer other research questions?
    A. Yes
        i. If yes, please provide specifics___________
    B. No
    C. Other, please specify___________________

16. Do you think that VIADS is a useful tool to facilitate research activities in general?
    A. Yes
        i. If yes, please provide specifics___________
    B. No
    C. Other, please specify___________________
