## Appendix A for "Scientific hypothesis generation process in clinical research: a secondary data analytic tool versus experience study protocol"

### Follow up survey at the end of hypothesis-generation tasks

1. Can you tell us your basic demographics:
    a. Gender: Female, Male
    b. Age range: <=35, 36-45, 46-55, 56-65, 66-75, >=76
    c. Job title:
    d. Type of organization that you affiliate with:

2. How often do you need to use computers?
    a. Daily
        i. <=2 hours
        ii. >2 and <=4 hours
        iii. >4 hours
    b. Weekly
    c. Monthly
    d. Yearly

3. How many years of experience do you have in clinical research (including hypothesis generation and study design)?
    a. <=2 years
    b. >2 and <5 years
    c. >=5 and <10 years
    d. >= 10 years

4. How many years of experience do you have in data analysis during clinical research?
    a. <=2 years
    b. >2 and <5 years
    c. >=5 and <10 years
    d. >= 10 years

5. What type of role do you mainly play in clinical research (including hypothesis generation and study design)?
    a. Leading role
    b. Participating role
    c. Other, please specify_______________

6. What type of role do you mainly play in data analysis during clinical research?
    a. Leading role
    b. Participating role
    c. Other, please specify_______________

7. What data analysis tools do you usually use? Please check all that apply.
    a. Excel
    b. R
    c. SAS
    d. SPSS
    e. Other, please specify____________________

8. In a 0 (the least helpful) to 10 (the most helpful) scale, what would be your assessment of the following dimensions utilized in formulating your hypothesis:

    a. Basic, applied, translational research;
    b. observational or experimental study;
    c. descriptive or analytic research;
    d. comparison, association, or causal study;
    e. retrospective or prospective research;
    f. longitudinal or cross-sectional research
    g. qualitative or quantitative research
    h. single variable or multiple variables
    i. variables: dependent, independent, moderator, control, and intervening
    j. focused population
    k. one-tailed or two-tailed
    l. investigational intervention
    m. outcomes of interest

9. Can you give examples of the attributes or instructions that you think would be very helpful in formulating a hypothesis, however, are not provided during the study session?

10. Do you have any additional comments or suggestions about how to facilitate hypothesis generation?
